# Missense variants in the voltage sensing and pore domain of *KCNH5* cause neurodevelopmental phenotypes including epilepsy

**DOI:** 10.1101/2022.04.26.22274147

**Authors:** Hannah C. Happ, Lynette G. Sadleir, Matthew Zemel, Guillem de Valles-Ibáñez, Michael S. Hildebrand, Allyn McConkie-Rosell, Marie McDonald, Halie May, Tristan Sands, Vimla Aggarwal, Christopher Elder, Timothy Feyma, Allan Bayat, Rikke S. Møller, Christina D. Fenger, Jens Erik Klint Nielsen, Anita N. Datta, Kathleen M. Gorman, Mary D. King, Natalia Linhares, Barbara K. Burton, Andrea Paras, Sian Ellard, Julia Rankin, Anju Shukla, Purvi Majethia, Rory J. Olson, Karthik Muthusamy, Lisa A Schimmenti, Keith Starnes, Lucie Sedláčková, Katalin Štěrbová, Markéta Vlčková, Petra Laššuthová, Alena Jahodová, Brenda E. Porter, Nathalie Couque, Estelle Colin, Clément Prouteau, Corinne Collet, Thomas Smol, Roseline Caumes, Fleur Vansenne, Francesca Bisulli, Laura Licchetta, Richard Person, Erin Torti, Kirsty McWalter, Richard Webster, Gaetan Lesca, Pierre Szepetowski, Ingrid E. Scheffer, Heather C. Mefford, Gemma L. Carvill

## Abstract

**Objective:** *KCNH5* encodes the voltage-gated potassium channel EAG2/Kv10.2. We aimed to delineate the neurodevelopmental and epilepsy phenotypic spectrum associated with *de novo KCNH5* variants.

**Methods:** We screened 893 individuals with developmental and epileptic encephalopathies (DEEs) for *KCNH5* variants using targeted or exome sequencing. Additional individuals with *KCNH5* variants were identified through an international collaboration. Clinical history, EEG, and imaging data were analyzed; seizure types and epilepsy syndromes were classified. We included three previously published individuals including additional phenotypic details.

**Results:** We report a cohort of 17 patients, including nine with a recurrent *de novo* missense variant p.Arg327His, four with a recurrent missense variant p.Arg333His, and four additional novel missense variants. All variants were located in or near the functionally critical voltage-sensing or pore domains, absent in the general population, and classified as pathogenic or likely pathogenic using American College of Medical Genetics and Genomics (ACMG) criteria. All individuals presented with epilepsy with a median seizure onset at six months. They had a wide range of seizure types, including focal and generalized seizures. Cognitive outcomes ranged from normal intellect to profound impairment. Individuals with the recurrent p.Arg333His variant had a self-limited drug-responsive focal or generalised epilepsy and normal intellect, while the recurrent p.Arg327His variant was associated with infantile-onset DEE. Two individuals with variants in the pore-domain were more severely affected, with neonatal-onset DEE, profound disability, and childhood death.

**Conclusions:** We report the first cohort of 17 individuals with pathogenic or likely pathogenic missense variants in the voltage sensing and pore domains of Kv10.2, including 14 previously unreported individuals. We present evidence for a putative emerging genotype-phenotype correlation with a spectrum of epilepsy and cognitive outcomes. Overall, we expand the role of EAG proteins in human disease and establish *KCNH5* as implicated in a spectrum of neurodevelopmental disorders (NDDs) and epilepsy.

## Introduction

The voltage-gated potassium channels are a large group of transmembrane proteins critical for controlling neuronal excitability and regulating electrophysiological properties in the brain. The ether-a-go-go (EAG) subfamily consists of two members, EAG1/Kv10.1 encoded by *KCNH1* and EAG2/Kv10.2 encoded by *KCNH5*. Pathogenic gain of function (GOF) variants in *KCNH1* are implicated in Temple-Baraitser and Zimmermann-Laband syndromes^1-3^; epilepsy is a common feature of these overlapping syndromes^4^. Individuals with variants in *KCNH5* have been reported in large exome sequencing cohort studies of individuals with neurodevelopmental disorders (NDDs). Of the three previously reported individuals, two had a recurrent missense variant (p.Arg327His) associated with a developmental and epileptic encephalopathy (DEE) with onset prior to 1 year of age ^5, 6^. The third had a neonatal-onset DEE and a different *de novo* missense variant (p.Ile463Thr)^7^. Despite these case reports, the *KCNH5*-associated phenotypes are not well delineated.

*KCNH5* is expressed in excitatory neurons in upper layer IV of the cerebral cortex and the hippocampus^8, 9^. *KCNH5* encodes for Kv10.2 which localizes to the somatodendritic region where it plays a role in controlling the electrical coupling between cell bodies and distal dendrites^9^. Like most potassium channels, it consists of four transmembrane domains (S1-S4) that make up the voltage-sensing domain, two transmembrane domains and a reentrant loop (S5-S6) that comprise the pore module (Fig. 1A, C). The recurrent Kv10.2 p.Arg327His variant localizes to S4, where voltage-clamp analysis in a heterologous expression system demonstrated a hyperpolarized shift of voltage dependence of activation and acceleration of activation, consistent with a gain-of-function pathogenic mechanism^10^.

**Figure 1.**
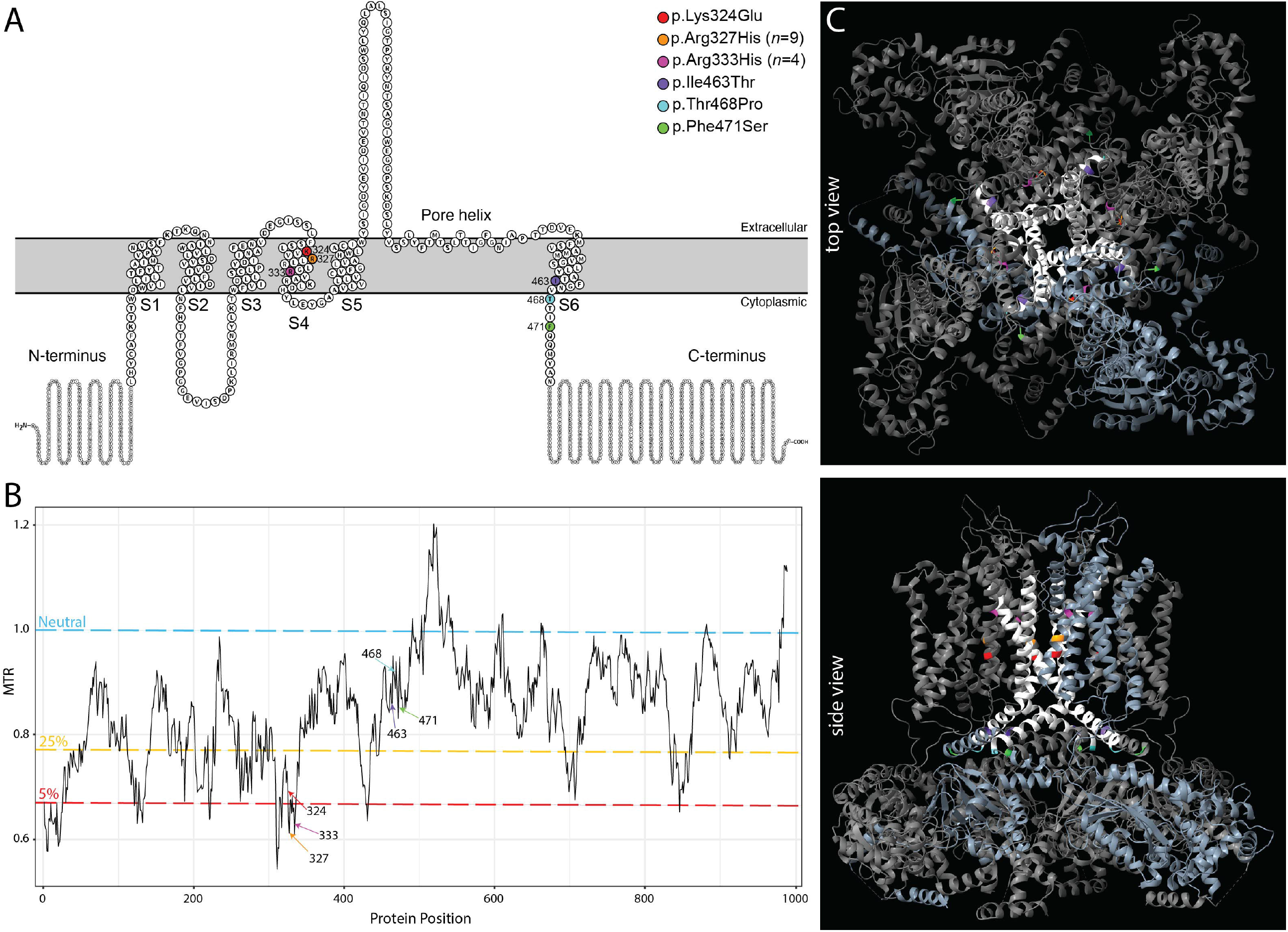
Location and conservation of the six unique *de novo KCNH5* missense variants. **A**. Schematic of Kv10.2 (EAG2), modified from Protter^29^ plot of Q8NCM2 (KCNH5_Human). Of the six transmembrane domains, S1-S4 make up the voltage sensing domain and S5-6 form the pore helix. The *KCNH5* patient-specific variants identified in this study are indicated by their amino acid change. **B**. Graph of missense tolerance ratio (MTR; y-axis) and protein position (x-axis), with variants indicated. MTR^30^ is a measure of protein-encoding cDNA sequence intolerance to missense variants. Variant positions with a value greater than one (blue line) is considered neutral; values below one are under constraint. Two variants (p.Arg327His, p.Arg333His) are in the 5^th^ percentile of least tolerated missense alterations in the exome. The 5^th^ and 25^th^ percentiles are highlighted in red and yellow, respectively. **C**. Locations of variants (color coded) are mapped onto the crystal structure of homotetrameric assembly of Kv10.1 (PDB5K7L); all variants are perfectly conserved between KCNH5 and KCNH1 and are mapped to the corresponding amino acid. The pore domain is highlighted in white, and a single tetrameric subunit is colored light blue.

Here we report a cohort of 17 individuals with *de novo* or inherited (n=1) missense variants in the voltage-sensing and pore domains of *KCNH5*. We describe 14 new patients and provide additional phenotypic information for the three published individuals from large cohort studies^5-7^. We analyze the spectrum of *KCNH5*-associated neurodevelopment disorders, particularly the epilepsy phenotypes, and discuss genotype-phenotype correlations.

## Methods

### Cohort

We performed *KCNH5* targeted resequencing in 680 individuals with DEEs and exome sequencing in 213 individuals with DEEs, totaling 893 individuals. All individuals underwent clinical assessment and medical records, EEG, and imaging data were obtained. Seizure types and epilepsy syndromes were classified according to the ILAE classification^11, 12^. Individuals with DEEs had been previously tested for pathogenic variants in the majority of known DEE genes (^13^ and unpublished data). The study was approved by the Austin Health Human Research Ethics Committee, the New Zealand Health and Disability Ethics committee and University Hospital of Lyon and Aix-Marseille University in France Ethics committee. Written informed consent including permission to publish was provided by the patient or their parent or legal guardian in the case of minors or those with intellectual disability.

We identified an additional 14 individuals with *KCNH5* variants through Genematcher^14^. These individuals were consented using research protocols approved by their local ethics committees. Their clinicians provided clinical information. For the three previously published individuals (probands 2, 10, and 15), we obtained additional clinical information. In total, the cohort consists of 19 individuals with *KCNH5* missense variants, with 17 having variants localized in or near the voltage-sensing or pore domains.

### KCNH5 variant identification and analysis

We used molecular inversion probes (MIPs) to capture all exons and five base pairs of flanking intronic *KCNH5* sequence; next generation sequencing and data analysis were performed as described previously^13^. We resequenced *KCNH5*, covering 95% of the gene at a depth of 50X (median across the cohort of 680 individuals). Exome sequencing (*n*=213) was performed via Epi25^15^. We considered only variants that were non-synonymous, altered the acceptor/donor splice sites, or indels that disrupted the coding frame. Only variants that were not present in gnomAD and TOPMed (see URLs) were considered for segregation analysis. The variants in the 14 individuals recruited by the matchmaker exchange network^14^ were identified via clinical or research genome (n=2), exome (n=10), or gene panel (n=2) sequencing ^15^. Library preparation and data analysis were performed using local protocols and computational pipelines. Variants were classified according to the American College of Medical Genetics and Genomics (ACMG) guidelines for variant classification^16^.

*KCNH1* (EAG1/Kv10.1) and *KCNH5* (EAG2/Kv10.2) have 72% identity at the amino acid level. The three-dimensional structure of Kv10.2 has not yet been resolved; thus, to model the location of each of the identified missense variants, we used the Kv10.2 homology model generated from the Kv10.1 cryo-EM structure (5k7l.1.A) using UCSF ChimeraX^17^.

## Results

### Rare missense variants in KCNH5

We identified two individuals with the previously reported *de novo* missense variant p.Arg327His in two DEE cohorts (1/680 by MIPs and 1/213 by exome^15^). Through our international collaboration, we identified five additional individuals with this recurrent variant, which arose *de novo* in four individuals (Table 1). We also identified five additional missense variants in the voltage-sensing and pore domains of Kv10.2 (Fig. 1). One variant (p.Arg333His) was present in four individuals; it arose *de novo* in three, and was inherited from an affected parent in the fourth proband. The four other *de novo* missense variants were each observed once in single individuals.

**Table 1.**
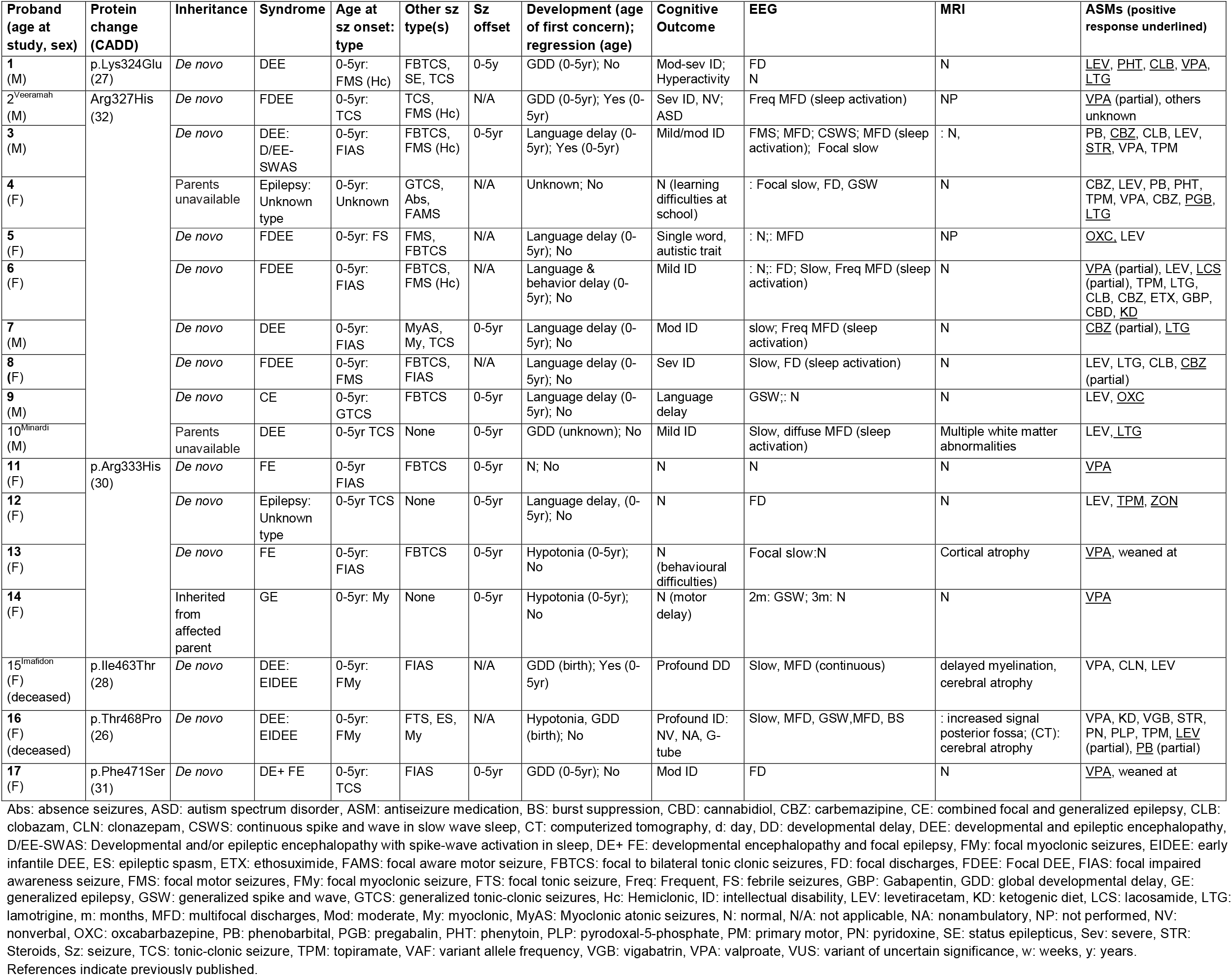
**Clinical presentation of our cohort and previously published individuals with P/LP *KCNH5* missense variants**.

The amino acids at all six variant positions (2 recurrent, 4 singletons) were conserved between Kv10 proteins. The missense variants had CADD^18^ scores of 26-32, conferring a high likelihood of deleteriousness (Fig 1B and Table 1). All variants were absent from the general population (gnomAD and TOPMed) and were classified as either pathogenic or likely pathogenic (P/LP) by ACMG criteria (eTable 1)^16^.

We also identified two *de novo* variants of uncertain significance (VUS) in the N-terminal (p.Leu181Pro) and C-terminal (p.Ile606Thr) of Kv10.2 in two individuals with NDDs without seizures (eAppendix). These VUS were located outside of the functionally critical pore or voltage-sensing domains. One variant (p.Leu181Pro) did not segregate with the NDD in the family, and the individual with the p.Ile606Thr variant also carried a pathogenic 16p11.2 microdeletion that may explain some or all of their clinical features. While these variants are included here for completeness, this study focuses on the 17 individuals with variants in’ the voltage-sensing or pore-forming domains.

### Phenotypic features in individuals with KCNH5 pathogenic or likely pathogenic variants

All 17 individuals with pathogenic or likely pathogenic missense variants in the voltage-sensing or pore domains had epilepsy (Table 1). Moreover, the parent of proband 14 who carries the recurrent p. Arg333His variant also has epilepsy, though detailed clinical information was not available. The median age of seizure onset was six months (range: day 1 to 12 months). A wide range of seizure types was seen: focal myoclonic seizures (2), focal impaired awareness seizures (FIAS) (8), focal to bilateral tonic-clonic seizures (FBTCS) (9), focal motor seizures (8), generalized myoclonic (2), generalized tonic-clonic seizure (GTCS) (2), myoclonic-atonic (1), absence seizure (1), unknown onset tonic-clonic seizures (6) and epileptic spasms (1). Nine (53%) individuals had a DEE. The remainder had focal epilepsy (3), generalized epilepsy (2) or there was inadequate information to make an epilepsy type diagnosis (3). Nine individuals (53%) achieved seizure freedom, with median seizure offset at 2.5 years (range: 3 months-8 years).

Developmental concerns were noted at a median age of 15 months in 14 individuals (range birth to 3.5 years), predominantly affecting language (11). One child had normal development and for two individuals early developmental information was limited. Cognitive outcome for the ten individuals over 5 years of age ranged from normal (3/10) to mild (3/10), moderate (2/10), severe (1/10) and profound (1/10) intellectual disability (ID).

For the nine individuals with the recurrent voltage-sensing domain p.Arg327His variant, seizure onset occurred at a median age of 6 months (range 2-10 months). For the eight individuals for whom an epilepsy syndrome diagnosis could be made, all but one, who was too young to assess, had a DEE (7/8), which was focal in five individuals. Proband 3 had the syndrome of epileptic encephalopathy with continuous spike-and-wave during sleep associated with language regression^11^. Sleep activation of epileptiform discharges was noted in 5 other patients but was not associated with developmental regression. The cognitive outcome for those over 5 years of age with this recurrent variant ranged from normal with learning difficulties (1/6) to mild (3/6), moderate (1/6), or severe ID (1/6). For those in whom the complete anti-seizure medication (ASM) history was available, their epilepsy was drug-resistant (8/9), with only 4/9 eventually becoming seizure-free.

The phenotype was much less severe for the four individuals with the second recurrent voltage-sensing domain p.Arg333His variant. This group had onset at a similar age (2-10 months) with either focal or generalized seizures. In contrast, their seizures were drug-responsive, particularly to valproate (3/4), and all became seizure free (aged 3 months to 5 years). Although 3 of the 4 individuals had developmental delay, they all had normal cognition.

Strikingly, probands 15 (p.Ile463Thr) and 16 (p.Thr468Pro), with missense variants in or at the junction of the S6 transmembrane pore-forming domain, had a much more severe phenotype. Both individuals had an early infantile DEE with neonatal-onset drug-resistant seizures, beginning on day one of life. Neuroimaging showed cortical atrophy. They had profound disability and premature death.

There were two additional individuals with likely pathogenic variants and epilepsy. Proband 1, with p.Lys324Glu variant in the voltage-sensing domain, had a similar phenotype to individuals with the nearby recurrent p.Arg327His variant. This individual had a focal DEE with focal seizures, developmental delay, evolving to moderate ID. Proband 17 had the p.Phe471Ser variant in the C-terminal cytoplasmic domain which was very close to the pore-forming domain. They had moderate ID, drug-responsive focal epilepsy and a hyperkinetic movement disorder. They did not have an epileptic encephalopathy and presented a similar epilepsy phenotype to patients with the recurrent p.Arg333His variant.

## Discussion

Potassium channel subunits are one of the most important groups of genes associated with epilepsy, in particular with the most severe group of epilepsies, the DEEs^19^. Here, we show that *de novo* pathogenic variants in *KCNH5* in 17 patients are frequently associated with a DEE but may also cause self-limited epilepsies. We describe the phenotypic spectrum, with particular emphasis on the phenotype-genotype correlations and the putative impact of these missense variants on Kv10.2 channel function.

We highlight a genotype-phenotype correlation for variants across the protein (Figure 2). The p.Arg333His variant is associated with the mildest epilepsy phenotype, a drug-responsive self-limited focal or generalized epilepsy with normal cognitive outcome. Conversely, the nearby most common recurrent p.Arg327His variant has a far more severe epilepsy phenotype of DEE. Interestingly, the nearby p.Lys324Glu voltage-sensing domain variant was also associated with a DEE. Individuals with these two variants predominantly presented with an infantile-onset DEE with drug-resistant generalized and focal seizures. Early developmental concerns were noted with cognitive outcome ranging from learning difficulties to severe ID. Finally, two individuals with variants in the pore-forming S6 transmembrane domain (p.Ile463Thr and p.Thr468Pro) had an even more severe phenotype of neonatal-onset DEE with onset on day one of life resulting in profound disability and death.

**Figure 2.**
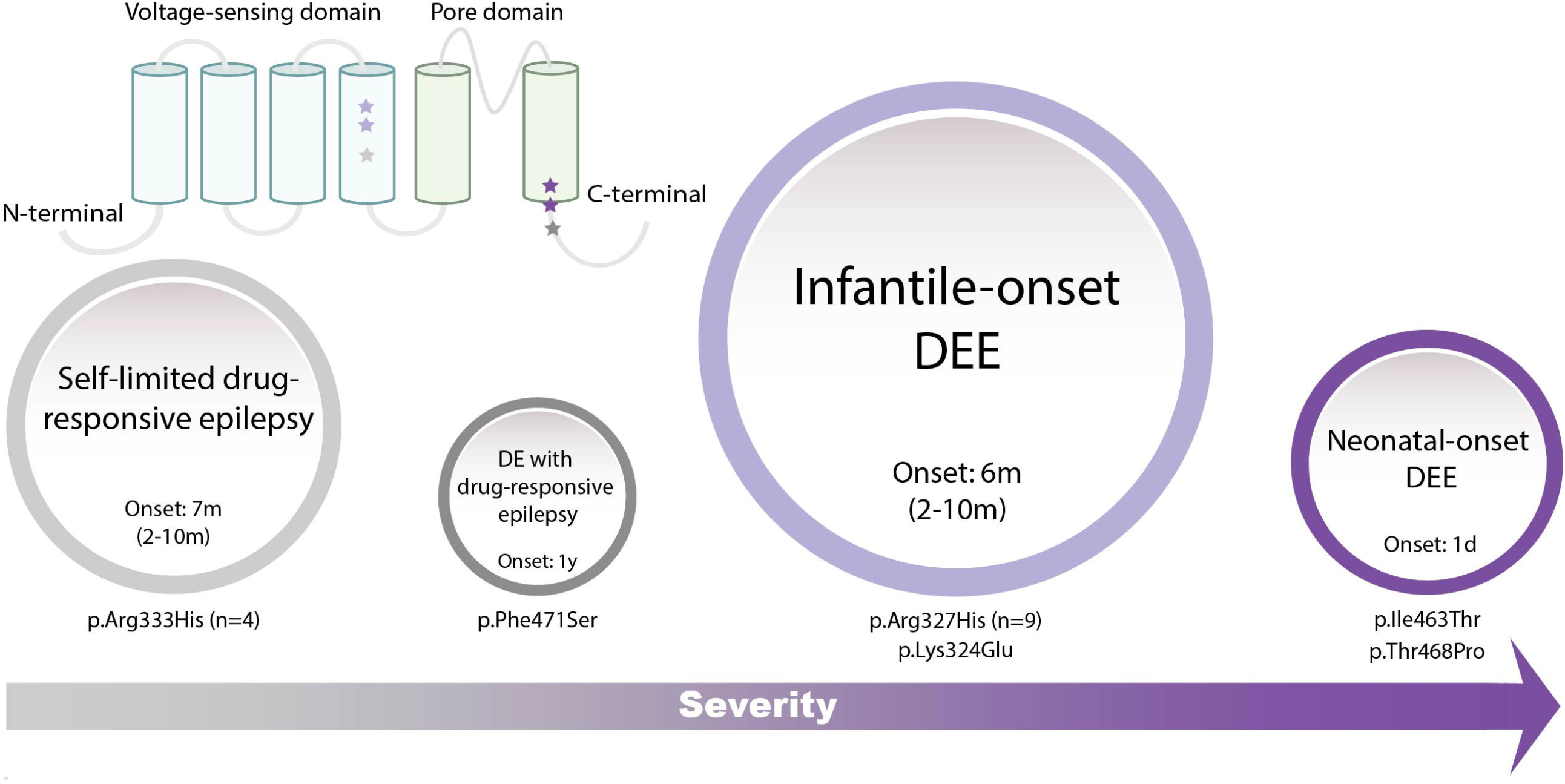
Phenotypic spectrum associated with *KCNH5* missense variants. The number of individuals in the phenotype group are represented by circle size and colors of each circle and star match the variant/phenotype class. The individuals with the p.Lys324Glu and p.Arg327His variants in the S4 transmembrane domain presented with an infantile-onset DEE with drug-resistant generalized and focal seizures, while the four individuals with the nearby recurrent p.Arg333His variant had drug-responsive seizures and became seizure-free. The two individuals with variants in or directly at the junction of the S6 transmembrane domain (p.Ile463Thr and p.468Pro) are the most severely affected with a neonatal-onset DEE. The single individual with the nearby p.Phe471Ser variant was less severely affected with moderate ID and drug-responsive seizures. Range is indicated in parentheses where applicable. DE: developmental encephalopathy, DEE: developmental and epileptic encephalopathy, m: month, d: day.

The p.Arg327His variant, located in the voltage-sensing S4 transmembrane domain, acts in a GOF manner^10^. Multistate structural modeling demonstrated that the p.Arg327His variant favors channel opening, and voltage-clamp experiments showed a hyperpolarized shift of voltage dependence of activation, as well as acceleration of activation^10^. These changes in electrophysiological properties are consistent with the GOF mechanism that is typically observed for several potassium channels implicated in DEEs, such as *KCNH1*^*1*^, *KCNT1*^*20*^, and *KCNT2*^*21*^. A subset of epilepsy-associated variants in *KCNA2, KCNB1, KCNQ2*, and *KCNQ3* have similarly been shown to act in a GOF manner^22-24^. How these changes lead to epilepsy remains unresolved^25^. We hypothesize that the new *KCNH5* voltage-sensing and pore domain pathogenic variants described here also have a similar GOF mechanism to the p.Arg327His variant. This is supported by *in silico* data including the tolerance of *KCNH5* to loss-of-function (truncating) variants in the general population. *KCNH5* has a probability of loss-of-function intolerant (pLI) of 0.02^26^. Epilepsy-associated genes that confer pathogenicity via haploinsufficiency typically have pLI scores ∼1, highlighting that loss of function is unlikely to be the pathogenic mechanism for *KCNH5* variants. Conversely, missense variants in the gene are not as well tolerated, with a z-score of 2.51 (Fig 1B), suggesting selection against certain missense variants throughout human evolution.

The two most severely affected individuals carried missense variants (p.Ile463Thr, p.Thr468Pro) that cluster right at the junction of the S6 transmembrane domain and the early C-terminal cytoplasmic domain that form the channel pore. The two missense variants are located within five amino acids of each other right at the channel pore. Missense variants are similarly found in and just outside the S6 domain of the EAG1/Kv10 pore encoded by *KCNH1* in individuals with Temple Baraitser syndrome, which includes epilepsy as a key feature. Detailed electrophysiological studies of these S6 missense variants in *KCNH1* showed delayed deactivation and decreased threshold of activation in both human cells and *Xenopus* oocytes^1^, consistent with a GOF mechanism. We hypothesize that the pore-forming *KCNH5* missense variants act in a similar GOF manner. Future studies are required to confirm this functional effect and determine whether missense pathogenic variants in the most central pore region have a more profound GOF effect on Kv10.2 function than those in the voltage-sensing domain or located further away from the pore.

*De novo* missense variants in *KCNH1*/Kv10.1 cause Temple-Baraitser syndrome (TBS) and Zimmermann-Laband syndrome (ZLS)^1-3^. Features of both include ID, epilepsy, facial dysmorphism, and nail hypoplasia^27^. GOF *KCNH1* missense variants have recently been reported in individuals with ID and epilepsy without the additional phenotypic features of TBS/ZLS^28^. Individuals with both syndromic and non-syndromic *KCNH1-*associated disorders with epilepsy have both focal and generalized seizures in infancy^4^. *KCNH1* and *KCNH5* are predominantly expressed in the adult central nervous system (Human Protein Atlas and GTeX, see URLs); however, current expression datasets are largely limited to adult tissues. Particularly as the variants described here are conserved between *KCNH1* and *KCNH5* (eFigure 1), future datasets such as the Developmental Genotype-Tissue Expression (dGTEx) initiative will be valuable for evaluating both the multi-system involvement of *KCNH1-* associated disorders and the CNS-limited *KCNH5*-related phenotypes.

Overall, we establish *KCNH5* as a gene associated with epilepsy and neurodevelopmental phenotypes and identify an emerging genotype-phenotype correlation. We describe 17 individuals with pathogenic or likely pathogenic variants in *KCNH5*, including four novel variants. The p.Arg327His variant located in the voltage-sensing domain causes gain of channel function^10^, and we hypothesize that the additional *KCNH5* voltage-sensing and pore domain variants also lead to epilepsy via a similar GOF mechanism. We highlight an intriguing genotype-phenotype correlation with a spectrum of epilepsy and cognitive outcomes. These observations will be further supported by examining the functional effects associated with the location of different variants in Kv10.2, with potential therapeutic implications. Our study expands the role of EAG proteins in human disease, highlighting that *KCNH5* variants are implicated in a spectrum of NDD and epilepsy phenotypes.

## Supporting information

strobe

## Data Availability

All data produced in the present work are contained in the manuscript

## Acknowledgements

We thank the patients and their families for participating in our research study. This work was supported by funding from the NIH (NINDS NS069605). H.C.M is a recipient of the Burroughs Wellcome Fund Career Award for Medical Scientists. G.L.C was funded by the Citizens United for Research in Epilepsy (CURE) Taking Flight Award and NIH (NINDS NS089858). I.E.S is supported by a National Health and Medical Research Council of Australia (NHMRC) Program grant, Investigator grant, Centre of Research Excellence and Synergy grant. L.G.S and G.V.I were supported by Health Research Council of New Zealand and Cure Kids research grants. L.S., K.S., M.V., P.L., A.J. are supported by the Ministry of Health of the Czech Republic AZV NU20-04-00279.

We thank the Epi25 principal investigators, local staff from individual cohorts, and all of the patients with epilepsy who participated in the study for making possible this global collaboration and resource to advance epilepsy genetics research. This work is part of the Centers for Common Disease Genomics (CCDG) program, funded by the National Human Genome Research Institute (NHGRI) and the National Heart, Lung, and Blood Institute (NHLBI). CCDG-funded Epi25 research activities at the Broad Institute, including genomic data generation in the Broad Genomics Platform, are supported by NHGRI grant UM1 HG008895 (PIs: Eric Lander, Stacey Gabriel, Mark Daly, Sekar Kathiresan). The Genome Sequencing Program efforts were also supported by NHGRI grant 5U01HG009088-02. The content is solely the responsibility of the authors and does not necessarily represent the official views of the National Institutes of Health. We thank the Stanley Center for Psychiatric Research at the Broad Institute for supporting the genomic data generation efforts.

## Ethical publication statement

We confirm that we have read the Journal’s position on issues involved in ethical publication and affirm that this report is consistent with those guidelines

## Accession numbers

*KCNH5* mRNA <u>**NM_139318.4**</u> and protein <u>**NP_647479.2**</u> sequence.

*KCNH1* protein <u>NP_758872.1</u>

## URLs

gnomAD: https://gnomad.broadinstitute.org

TopMED: https://bravo.sph.umich.edu/freeze3a/hg19/

Human Protein Atlas: https://www.proteinatlas.org/

GTEx: https://www.gtexportal.org/home/

## References

1. Simons C, Rash LD, Crawford J, et al. Mutations in the voltage-gated potassium channel gene KCNH1 cause Temple-Baraitser syndrome and epilepsy. Nat Genet. United States 2015: 73–77.

2. Kortum F, Caputo V, Bauer CK, et al. Mutations in KCNH1 and ATP6V1B2 cause Zimmermann-Laband syndrome. Nat Genet 2015;47:661–667.

3. Gripp KW, Smithson SF, Scurr IJ, et al. Syndromic disorders caused by gain-of-function variants in KCNH1, KCNK4, and KCNN3-a subgroup of K(+) channelopathies. Eur J Hum Genet 2021;29:1384–1395.

4. Mastrangelo M, Scheffer IE, Bramswig NC, et al. Epilepsy in KCNH1-related syndromes. Epileptic Disord 2016;18:123–136.

5. Veeramah KR, Johnstone L, Karafet TM, et al. Exome sequencing reveals new causal mutations in children with epileptic encephalopathies. Epilepsia 2013;54:1270–1281.

6. Minardi R, Licchetta L, Baroni MC, et al. Whole-exome sequencing in adult patients with developmental and epileptic encephalopathy: It is never too late. Clin Genet 2020;98:477–485.

7. Imafidon ME, Sikkema-Raddatz B, Abbott KM, et al. Strategies in Rapid Genetic Diagnostics of Critically Ill Children: Experiences From a Dutch University Hospital. Front Pediatr 2021;9:600556.

8. Saganich MJ, Vega-Saenz de Miera E, Nadal MS, Baker H, Coetzee WA, Rudy B. Cloning of components of a novel subthreshold-activating K(+) channel with a unique pattern of expression in the cerebral cortex. The Journal of neuroscience: the official journal of the Society for Neuroscience 1999;19:10789–10802.

9. Jeng CJ, Chang CC, Tang CY. Differential localization of rat Eag1 and Eag2 K+ channels in hippocampal neurons. Neuroreport 2005;16:229–233.

10. Yang Y, Vasylyev DV, Dib-Hajj F, et al. Multistate structural modeling and voltage-clamp analysis of epilepsy/autism mutation Kv10.2-R327H demonstrate the role of this residue in stabilizing the channel closed state. The Journal of neuroscience: the official journal of the Society for Neuroscience. United States 2013: 16586–16593.

11. Scheffer IE, Berkovic S, Capovilla G, et al. ILAE classification of the epilepsies: Position paper of the ILAE Commission for Classification and Terminology. Epilepsia 2017;58:512–521.

12. Fisher RS, Cross JH, French JA, et al. Operational classification of seizure types by the International League Against Epilepsy: Position Paper of the ILAE Commission for Classification and Terminology. Epilepsia 2017;58:522–530.

13. Carvill GL, Heavin SB, Yendle SC, et al. Targeted resequencing in epileptic encephalopathies identifies de novo mutations in CHD2 and SYNGAP1. Nat Genet 2013;45:825–830.

14. Sobreira N, Schiettecatte F, Valle D, Hamosh A. GeneMatcher: a matching tool for connecting investigators with an interest in the same gene. Human mutation 2015;36:928–930.

15. Epi25 Collaborative. Electronic address sbuea, Epi C. Ultra-Rare Genetic Variation in the Epilepsies: A Whole-Exome Sequencing Study of 17,606 Individuals. Am J Hum Genet 2019;105:267–282.

16. Richards S, Aziz N, Bale S, et al. Standards and guidelines for the interpretation of sequence variants: a joint consensus recommendation of the American College of Medical Genetics and Genomics and the Association for Molecular Pathology. Genetics in medicine: official journal of the American College of Medical Genetics 2015;17:405–424.

17. Pettersen EF, Goddard TD, Huang CC, et al. UCSF ChimeraX: Structure visualization for researchers, educators, and developers. Protein Sci 2021;30:70–82.

18. Rentzsch P, Schubach M, Shendure J, Kircher M. CADD-Splice-improving genome-wide variant effect prediction using deep learning-derived splice scores. Genome Med 2021;13:31.

19. McTague A, Howell KB, Cross JH, Kurian MA, Scheffer IE. The genetic landscape of the epileptic encephalopathies of infancy and childhood. Lancet Neurol 2016;15:304–316.

20. Barcia G, Fleming MR, Deligniere A, et al. De novo gain-of-function KCNT1 channel mutations cause malignant migrating partial seizures of infancy. Nat Genet 2012;44:1255–1259.

21. Ambrosino P, Soldovieri MV, Bast T, et al. De novo gain-of-function variants in KCNT2 as a novel cause of developmental and epileptic encephalopathy. Ann Neurol 2018;83:1198–1204.

22. Miceli F, Soldovieri MV, Ambrosino P, et al. Early-onset epileptic encephalopathy caused by gain-of-function mutations in the voltage sensor of Kv7.2 and Kv7.3 potassium channel subunits. The Journal of neuroscience: the official journal of the Society for Neuroscience 2015;35:3782–3793.

23. Torkamani A, Bersell K, Jorge BS, et al. De novo KCNB1 mutations in epileptic encephalopathy. Ann Neurol 2014;76:529–540.

24. Syrbe S, Hedrich UBS, Riesch E, et al. De novo loss-or gain-of-function mutations in KCNA2 cause epileptic encephalopathy. Nat Genet 2015;47:393–399.

25. Niday Z, Tzingounis AV. Potassium Channel Gain of Function in Epilepsy: An Unresolved Paradox. Neuroscientist 2018;24:368–380.

26. Karczewski KJ, Francioli LC, Tiao G, et al. The mutational constraint spectrum quantified from variation in 141,456 humans. Nature 2020;581:434–443.

27. Hamilton MJ, Suri M. “Electrifying dysmorphology”: Potassium channelopathies causing dysmorphic syndromes. Adv Genet 2020;105:137–174.

28. Aubert Mucca M, Patat O, Whalen S, et al. Patients with KCNH1-related intellectual disability without distinctive features of Zimmermann-Laband/Temple-Baraitser syndrome. J Med Genet 2021.

29. Omasits U, Ahrens CH, Muller S, Wollscheid B. Protter: interactive protein feature visualization and integration with experimental proteomic data. Bioinformatics 2014;30:884–886.

30. Silk M, Petrovski S, Ascher DB. MTR-Viewer: identifying regions within genes under purifying selection. Nucleic Acids Res 2019;47:W121–W126.

