## Supplementary material for "Missense variants in the voltage sensing and pore domain of *KCNH5* cause neurodevelopmental phenotypes including epilepsy": Table e1

**Authors and affiliations**

Hannah C. Happ^1^*, Lynette G. Sadleir^2^*, Matthew Zemel^3^, Guillem de Valles-Ibáñez^2^, Michael S. Hildebrand^4^, Allyn McConkie-Rosell^6^, Marie McDonald^6^, Halie May^7^, Tristan Sands^7^, Vimla Aggarwal^8^, Christopher Elder^9^, Timothy Feyma^10^, Allan Bayat^11,12^, Rikke S. Møller^11,12^, Christina D. Fenger^11,13^, Jens Erik Klint Nielsen^14^, Anita N. Datta^15^, Kathleen M. Gorman^16,17^, Mary D. King^16,17^, Natalia Linhares^18^, Barbara K. Burton^19,20^, Andrea Paras^19,20^, Sian Ellard^21,22^, Julia Rankin^23^, Anju Shukla^24^, Purvi Majethia^24^, Rory J. Olson^25^, Karthik Muthusamy^25,26^, Lisa A Schimmenti^25,26^, Keith Starnes^27^, Lucie Sedláčková^28,31^, Katalin Štěrbová^29,31^, Markéta Vlčková^30,31^, Petra Laššuthová^28,31^, Alena Jahodová^29,31^, Brenda E. Porter^32^, Nathalie Couque^33^, Estelle Colin^34^, Clément Prouteau^34^, Corinne Collet^33^, Thomas Smol^35^, Roseline Caumes^36^, Fleur Vansenne^37^, Francesca Bisulli^38,39^, Laura Licchetta^39^, Richard Person^40^, Erin Torti^40^, Kirsty McWalter^40^, Richard Webster^4,41^, Gaetan Lesca^42^, Pierre Szepetowski^43^, Ingrid E. Scheffer^4,44,45^, Heather C. Mefford^46^**, Gemma L. Carvill^1,20,47^**

^1^Ken and Ruth Davee Department of Neurology, Northwestern University Feinberg School of Medicine, Chicago, IL, USA

^2^University of Otago, Wellington, New Zealand

^3^University of Washington, Seattle, WA, USA

^4^Epilepsy Research Centre, Department of Medicine, The University of Melbourne, Austin Health, Heidelberg, Victoria, Australia

^6^Duke University Medical Center, Durham, NC, USA

^7^Institute for Genomic Medicine, Columbia University Irving Medical Center, New York, NY, USA

^8^Department of Pathology and Cell Biology, Columbia University Irving Medical Center, New York, NY, USA

^9^Department of Neurology, Columbia University Irving Medical Center, New York, NY, USA

^10^Gillette Children's Specialty Healthcare, St. Paul, MN, USA

^11^Department of Epilepsy Genetics and Personalized Medicine, Danish Epilepsy Center, Dianalund, Denmark

^12^Institute of Regional Health Research, University of Southern Denmark, Denmark

^13^Amplexa Genetics, Odense, Denmark

^14^Department of Clinical Medicine, Zealand University Hospital, Roskilde, Denmark

^15^University of British Columbia, Vancouver, Canada

^16^The Department of Neurology and Clinical Neurophysiology, Children’s Health Ireland at Temple St., Temple Street, Dublin 1, Ireland

^17^School of Medicine and Medical Science, University College Dublin, Dublin 4, Ireland

^18^Genuity Science, Dublin, Ireland

^19^Ann & Robert H. Lurie Children’s Hospital of Chicago, Chicago, IL, USA

^20^Department of Pediatrics, Northwestern University Feinberg School of Medicine, Chicago, IL, USA

^21^Exeter Genomics Laboratory, Royal Devon University Healthcare NHS Foundation Trust, Exeter, UK

^22^Institute of Clinical and Biomedical Science, University of Exeter, UK

^23^Dept Clinical Genetics, Royal Devon University Healthcare NHS Foundation Trust, Exeter, UK

^24^Department of Medical Genetics, Kasturba Medical College, Manipal, Manipal Academy of Higher Education, Manipal, India

^25^Center for Individualized Medicine, Mayo Clinic, Rochester, MN, USA

^26^Department of Clinical Genomics, Mayo Clinic, Rochester, MN, USA

^27^Department of Neurology, Mayo Clinic, Rochester, MN, USA

^28^Neurogenetic Laboratory, Department of Pediatric Neurology, Second Faculty of Medicine, Charles University in Prague and Motol University Hospital, Prague, Czech Republic

^29^Department of Pediatric Neurology, Second Faculty of Medicine, Charles University in Prague and Motol University Hospital, Prague, Czech Republic

^30^Biology and Medical Genetics, Second Faculty of Medicine, Charles University in Prague and Motol University Hospital, Prague, Czech Republic

^31^Epilepsy Research Centre Prague – EpiReC consortium; Motol University Hospital is a full member of the ERN EpiCARE

^32^Stanford University School of Medicine, Palo Alto, CA, USA

^33^Laboratoire de biologie médicale multisites Seqoia-FMG2025, Laboratoire Génétique Moléculaire Robert-Debré, Paris, France

^34^Service de Génétique, CHU d'Angers, Angers, France

^35^Univ. Lille, CHU Lille, ULR7364 – RADEME, Institut de Genetique Medicale, F-59000 Lille, France

^36^Univ. Lille, CHU Lille, ULR7364 – RADEME, Clinique de Genetique, F-59000 Lille, France

^37^Univeristy Medical Center Groningen, Groningen, the Netherlands

^38^Department of Biomedical and NeuroMotor Sciences, University of Bologna, Bologna, Italy

^39^IRCCS Istituto delle Scienze Neurologiche di Bologna, Full Member of the ERN EpiCARE Bologna, Italy

^40^GeneDx, Gaithersburg, MD, USA

^41^T.Y. Nelson Department of Neurology and Neurosurgery, Children's Hospital at Westmead, Westmead, New South Wales, Australia

^42^ Department of Medical Genetics, University Hospital of Lyon, Claude Bernard Lyon 1 University, Lyon, France

^43^INSERM, Aix-Marseille University, INMED, Marseille, France

^44^Department of Neurology, Royal Children’s Hospital, Department of Paediatrics, The University of Melbourne, and Murdoch Children’s Research Institute, Parkville, Victoria, Australia

^45^The Florey Institute of Neuroscience and Mental Health, Victoria, Australia

^46^St. Jude Children's Research Hospital, Memphis, TN, USA

^47^Department of Pharmacology, Northwestern University Feinberg School of Medicine, Chicago, IL, USA

*These authors contributed equally to this manuscript

**These authors contributed equally to this manuscript

**eTable 1. Genetic details and additional findings of individuals with P/LP *KCNH5* missense variants.**

| **Proband** | **Variant** | **NGS** | **Additional findings** | **ACMG criteria** | **ACMG classification** |
| --- | --- | --- | --- | --- | --- |
| **1** | Chr14:g.63417250T>C  c.970A>G  p.Lys324Glu | Exome | None | PS2_Moderate, PM2, PP2, PP3 | Likely pathogenic |
| **2^Veeramah^** | Chr14:g.63417240C>T  c.980G>A  p.Arg327His | Exome | None | PS2_Very strong, PS3_Moderate, PM2, PP2, PP3 | Pathogenic |
| **3** |  | MIPs | None |  |  |
| **4** |  | Exome | None |  |  |
| **5** |  | Panel | None |  |  |
| **6** |  | Exome | None |  |  |
| **7** |  | Exome | None |  |  |
| **8** |  | Exome | None |  |  |
| **9** |  | Exome | None |  |  |
| **10^Mindardi^** |  | Exome | None |  |  |
| **11** | Chr14:g.63417222C>T  c.998G>A  p.Arg333His | Exome | None | PS2, PM2, PP2, PP3 | Likely pathogenic |
| **12** |  | Panel | None |  |  |
| **13** |  | Genome | None |  |  |
| **14** |  | Exome | None |  |  |
| **15^Imafidon^** | Chr14:g.63316552A>G  c.1388T>C  p.Ile463Thr | Exome | *de novo* VUS in *WDR47* (chr1:g.109524364G>C; c.2392C>G; p.Arg798Gly; not present in gnomAD) | PS2_Moderate, PM2, PP2, PP3 | Likely pathogenic |
| **16** | Chr14:g.63316538T>G  c.1402A>C  p.Thr468Pro | Exome | None | PS2_Moderate, PM2, PP2, PP3 | Likely pathogenic |
| **17** | Chr14:g.63316528A>G  c.1412T>C  p.Phe471Ser | Genome | None | PS2_Moderate, PM2, PP2, PP3 | Likely pathogenic |

MIPs: molecular inversion probes, VUS: variant of uncertain significance.

References indicate previously published. Please note the p.I463T variant was previously reported as a VUS.

**eAppendix 1**

The following VUS were identified in individuals with NDDs without seizures.

**Chr14:g.63453797A>G; c.542T>C; p.L181P** (CADD: 28)

This *de novo* variant in the N-terminal domain of Kv10.2 was detected by exome sequencing in a male who presented with mild fine motor and language delay. This individual went on to have learning difficulties, ASD, oppositional defiant disorder, and attention-deficit-hyperactivity disorder. An MRI showed a Chiari I malformation and spinal cord syrinx. This individual also carries a inherited VUS in *KCNH5* (chr14:g.63417122T>C; c.1098A>G; p.I366M; gnomAD VAF 0.001). He has a sibling with a similar but more severe phenotype who also carries the inherited VUS but who does NOT carry the *de novo* p.L181P variant, thus the variant does not segregate with the phenotype in this family.

**Chr14:g.63269052A>G; c.1817T>C; p.I606T** (CADD: 27)

This *de novo* variant in the C-terminus of Kv10.2 was detected by exome sequencing in a male who presented with global developmental delay who now has limited language and ASD. An EEG was normal. This individual also carries a inherited 16p12.2 microdeletion, which has been previously associated with various neurodevelopmental disorders (NDDs), including developmental delay^1^.

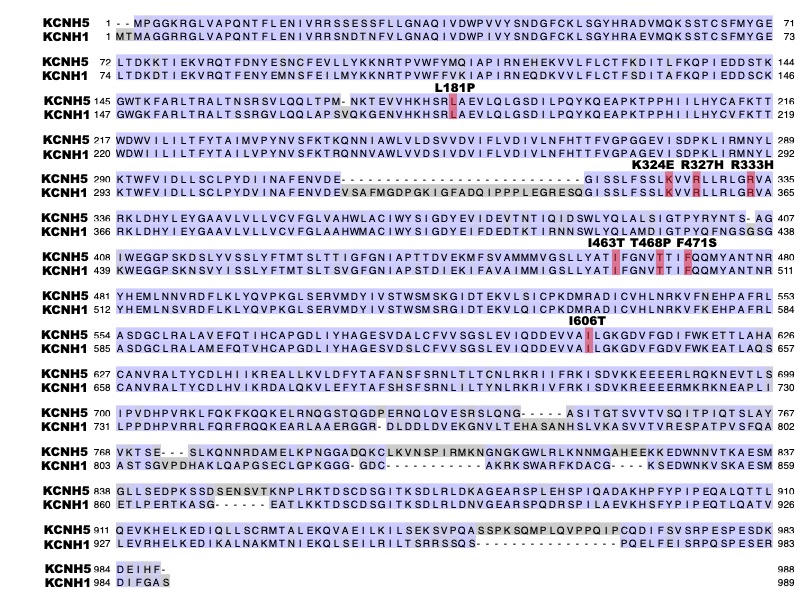

**eFigure 1. Conservation of the six missense variants between *KCNH1* and *KCNH5*.** *KCNH1* (EAG1/Kv10.1) and *KCNH5* (EAG2/Kv10.2) have 72% identity at the amino acid level. All NDD-associated missense variants (red highlight) are perfectly conserved between *KCNH1* and *KCNH5* at both the gene and amino acid level.

**eReferences**

1.	Girirajan S, Pizzo L, Moeschler J, Rosenfeld J. 16p12.2 Recurrent Deletion. In: Adam MP, Ardinger HH, Pagon RA, et al., eds. GeneReviews((R)). Seattle (WA)1993.
